# Implications of climatic and demographic change for seasonal influenza dynamics and evolution

**DOI:** 10.1101/2021.02.11.21251601

**Authors:** Rachel E. Baker, Qiqi Yang, Colin J. Worby, Wenchang Yang, Chadi M. Saad-Roy, Cecile Viboud, Jeffrey Shaman, C. Jessica E. Metcalf, Gabriel Vecchi, Bryan T. Grenfell

## Abstract

Seasonal influenza causes a substantial public health burden, as well as being a key substrate for pandemic emergence. Future climatic and demographic changes may alter both the magnitude, frequency and timing of influenza epidemics and the prospects for pathogen evolution, however, these issues have not been addressed systematically. Here, we use a parsimonious influenza model, grounded in theoretical understanding of the link between climate, demography and transmission to project future changes globally. We find that climate change generally acts to reduce the intensity of influenza epidemics as specific humidity increases. However, this reduction in intensity is accompanied by increased seasonal epidemic persistence with latitude, which may increase suitability for year-round local influenza evolution. Using a range of population growth scenarios, we find that the number of global locations with high evolution suitability may double by 2050. High population growth in tropical Africa could thus make this region a locus of novel strain emergence, shifting the current focus from South East Asia.

## Introduction

Climatic and demographic change will alter the landscape of infectious disease burden over the coming century [1, 2]. Characterizing these changes is a key public health challenge, and will determine our ability to predict and respond to future outbreaks. While advances have been made in characterizing the future of vector-borne diseases [3, 4, 5], less attention has been paid to directly-transmitted infections, despite evidence that climate and demography play a crucial role in determining the timing and intensity of these types of outbreaks [6, 7, 8]. Both laboratory and observational studies have uncovered a role for climate in driving influenza transmission [9, 10]. Demographic factors, such as population size and the recruitment of new susceptibles, are of great importance for driving disease dynamics, particularly for immunizing infections [8, 11, 12]. The high mortality burden from seasonal influenza, and lack of a consistently effective vaccine in the face of immune escape, makes characterizing future changes an important task [13, 14, 15]. Pandemic influenza is also a major potential threat, but complex drivers make potential climatic and demographic links elusive.

Influenza cases display distinct seasonal cycles, depending on latitude. In temperate locations at northern and southern latitudes, influenza epidemics are strongly seasonal, with high numbers of cases in the winter months and close to zero cases in the summer months [16, 8]. In tropical locations, epidemic patterns are less distinct and cases persist throughout the year, though higher case numbers have been observed during periods of elevated rainfall [16, 17, 18, 19]. Specific humidity has been to shown to play a role in driving these seasonal dynamics [20, 10, 21, 9]. Low specific humidity is correlated with increased influenza transmission and virus survival [20, 22], and has been shown to increase transmission in laboratory settings [21, 9, 20]. Large seasonal fluctuations in specific humidity can explain the intensity of outbreaks in temperate regions [10, 8]. More moderate variability in humidity, closer to the equator, can explain the less distinct influenza seasonality in tropical regions [22, 16].

A key challenge for influenza control is that gradual evolution of the virus’s surface glycoproteins (hemagglutinin and neuraminidase) leads to immune escape, a process known as antigenic drift [13]. This allows the virus to evade prevailing host population immunity, contributing to the high disease burden of influenza and the difficulty in providing a comprehensive vaccine [13, 14]. The population-level dynamics of seasonal influenza may influence the evolution of the influenza virus [23]. In temperate locations, a virus population bottleneck driven by the lack of cases in the summer months seasonally limits this local evolution [19, 14, 24]. In contrast, the continual circulation of the virus in populous tropical locations is expected to play an important role in the emergence of novel strains [25, 19, 16, 26, 27, 28, 14]. High and persistent viral abundance is theoretically linked to a higher population mutation rate, and therefore increased genetic diversity [29], as well as allowing for the importation and propagation of novel strains from other locations [27, 24].

Here we leverage a humidity-dependent influenza model, based on laboratory and observational studies [20], to predict population-level seasonal influenza dynamics globally. Using this model, we consider how changes to the climate will alter future influenza dynamics and how these changing dynamics may influence evolution. We project future influenza changes by coupling the influenza model to both highly resolved climate data from the present, and future climate changes projected by the Coupled Model Intercomparison Project phase 5 (CMIP5). We also consider how changes to future populations may affect the absolute depth of incidence troughs. Using population projections, in concert with climate change simulations, we evaluate future patterns of evolution propensity. Capturing the full dynamics of seasonal influenza evolution is a formidable task with many remaining uncertainties [30, 31, 32]; we therefore adopt a more synoptic approach. Population genetics theory suggests we can approximate the propensity for local evolution in terms of local persistence through the seasonal cycle i.e. the depth of incidence troughs [29, 33]. Our metric for year-round evolution suitability is *harmonic mean incidence* (HMI), which is correlated with trough depth, and shown to be related to pathogen genetic diversity [29, 34, 35, 36, 33] (Fig. 2, Fig. S2, see Methods).

## Results

Greenhouse gas induced climate change will lead to an increase in global temperatures and a corresponding increase of mean specific humidity (Fig. 1a). Specific humidity and influenza transmission have an inverse relationship in our model [10, 20] (Fig. S1). As specific humidity increases the basic reproductive number *R*_0_ (a measure of transmission defined as the number of secondary cases from a primary infection in a completely susceptible population) is expected to decline slightly (Fig. 1b). These declines will be non-uniform throughout the year such that the seasonal range of effective reproductive numbers will increase in some locations and decline in others (Fig. 1c).

**Figure 1:**
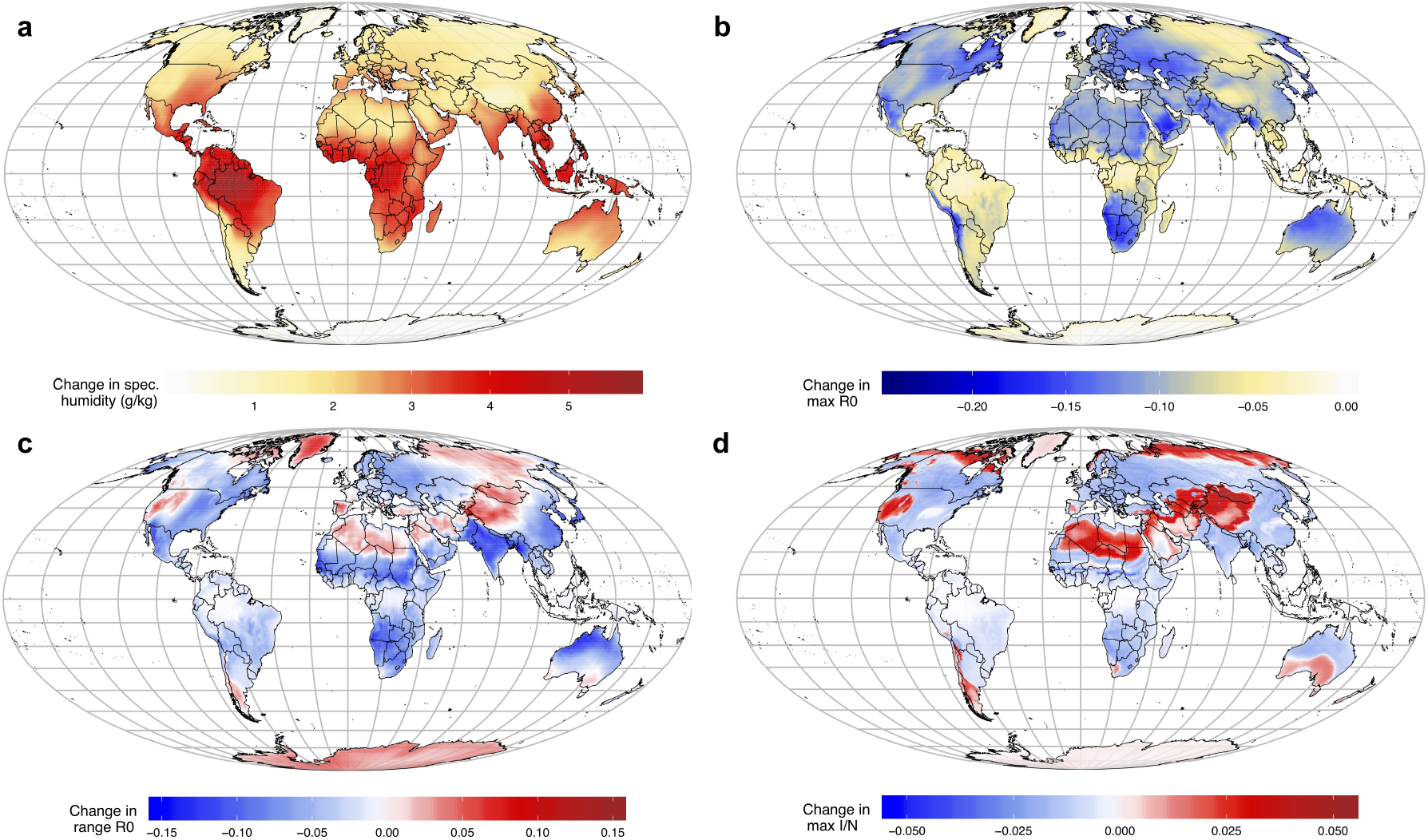
Climate change and influenza transmission. Projected 2010-2100 changes, based on CMIP5 multi-model mean under the RCP8.5. scenario, to a) annual mean specific humidity b) annual maximum *R*_0_ c) seasonal range in *R*_0_ d) seasonal max incidence per population (proportion of individuals in the infected category).

We run our influenza model for each location in our dataset using the projected transmission rates, setting population to one so that our results are the proportion of individuals in the susceptible, infected and recovered class. Fig 1d shows the change in peak annual incidence in our 2100 simulation versus 2010 simulation. As expected from a general humidity increase, in most locations peak incidence declines, meaning that climate change will lead to less intense outbreaks of influenza in the future. For some locations, we see an increase in intensity, though this tends to happen in less populous areas, such as desert locations (we address population distributions explicitly below).

These changes to dynamics may affect the evolution of the influenza virus. Harmonic mean incidence (HMI), a measure of the extent to which incidence persists throughout the year, is expected to increase with climate change in tropical locations (as detailed below, Fig. 3). Harmonic mean incidence can be used to approximate the population bottleneck that seasonal influenza virus faces in locations where cases go to zero during certain months. Places with very low (close to zero) HMI will experience a bottleneck, whereas places with high HMI will have more opportunities for year-round local evolution.

We test this hypothesis using phylogenetic data from six global locations for influenza A subtype H1NX and influenza A subtype H3NX. We chose locations across a range of tropical and temperate climates where there is sufficient data to estimate nucleotide diversity. Despite this, the data is likely biased by different sampling procedures across locations and time points. As such, we can only draw qualitative conclusions from the strain data.

Fig. 2a shows the nucleotide diversity for H1NX and H3NX subtypes in six locations, averaged by season (three-month period, starting in January). Broadly, we observe that locations with a more tropical climate, such as Hawaii, tend to experience year-round higher diversity of strains. Locations in more temperate regions, such as England, tend to experience lower year-round diversity. However, there are subtle effects across subtype and season. New York, a temperate location, experiences relative high diversity of H3NX strains in January-March, when case numbers are highest, with diversity declining in the later summer. Fig. 2b shows the minimum seasonal nucleotide diversity plotted against modeled harmonic mean incidence. We use modeled HMI* (* denotes per capita), as opposed to observed HMI*, because different definitions are used to record influenza cases across locations (for example, Taiwan records severe complicated influenza and the US records influenza-like illnesses). Using the HMI* predicted by our model provides both a common definition, while allowing us to investigate the predictive power of our approach. We find a significant positive relationship between modeled HMI* and minimum nucleotide diversity for H1NX (*p* = 0.029) and a positive, non-significant relationship for H3NX (*p* = 0.320) (though we have very limited data points in both cases). The relationship is stronger for H1NX diversity, where Hawaii appears to have approximately 4 times the diversity of New York. The relationship is weaker for H3NX where Hawaii has approximately 1.6 times the diversity of New York.

**Figure 2:**
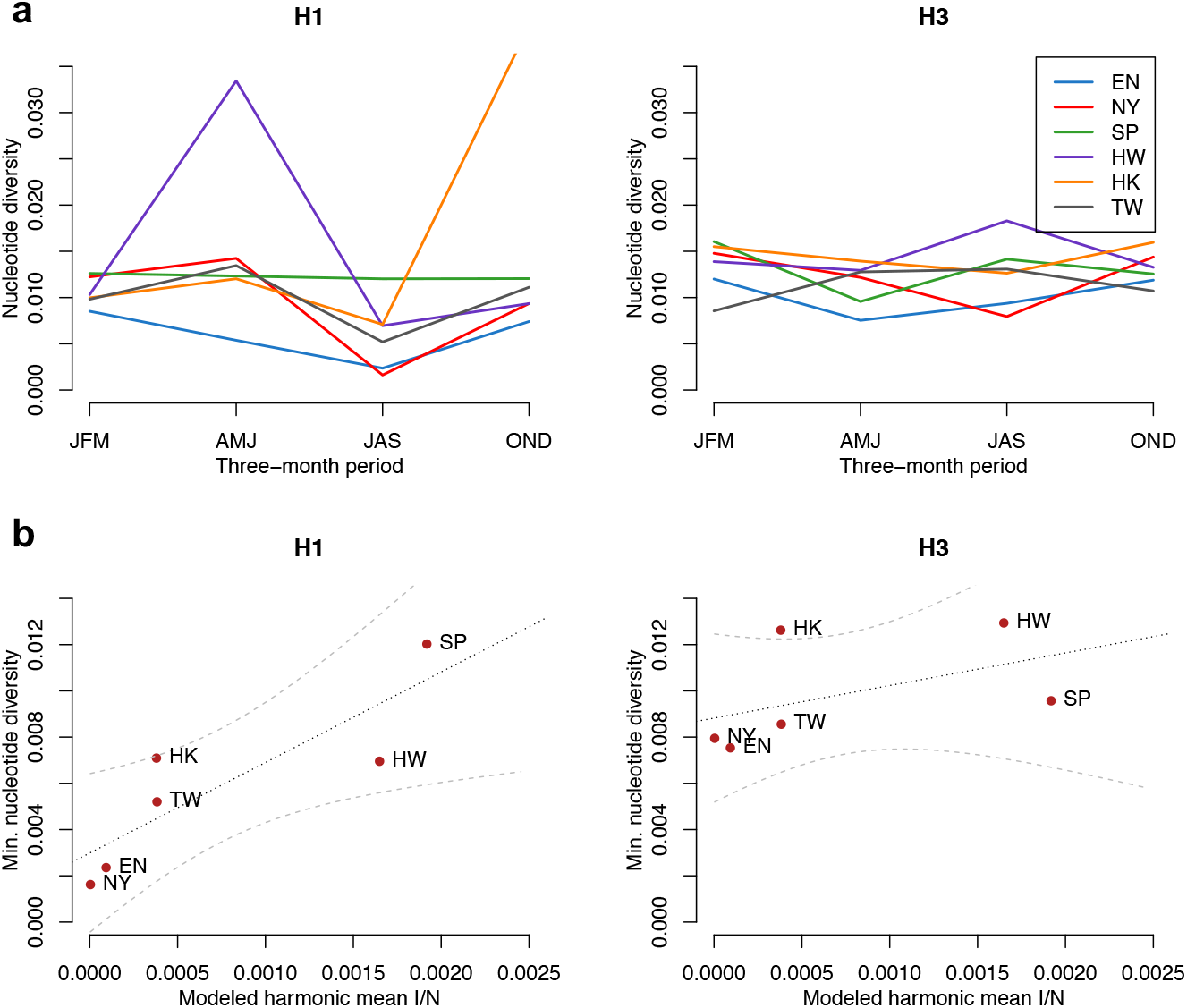
Nucleotide diversity and HMI. Average nucleotide diversity for six locations (England (EN), New York state (NY), Singapore (SP), Hawaii (HW), Hong Kong (HK) and Taiwan (TW)) for influenza subtypes H1NX and H3NX. Nucleotide diversity is plotted by season in a). Seasonal minimum nucleotide diversity is plotted against modelled harmonic mean incidence in b).

Fig 3a shows the global change in HMI* between 2010 and 2100 due to climate change. An increase in HMI* is visible as an expansion around the tropics. Fig 3b shows model projections for different example locations. High latitude locations, such as London, see minimal changes to HMI*, whereas mid-latitude locations, such as Hyderabad, see larger changes. Based on our fitted model, the change in HMI* in Hyderabad between 2010 and 2100 would lead to a 47% increase in minimum nucleotide diversity based on H1NX and a 8% increase based on H3NX. Similarly, Miami would be see a predicted 24% (H1) and 6% (H3) increase.

**Figure 3:**
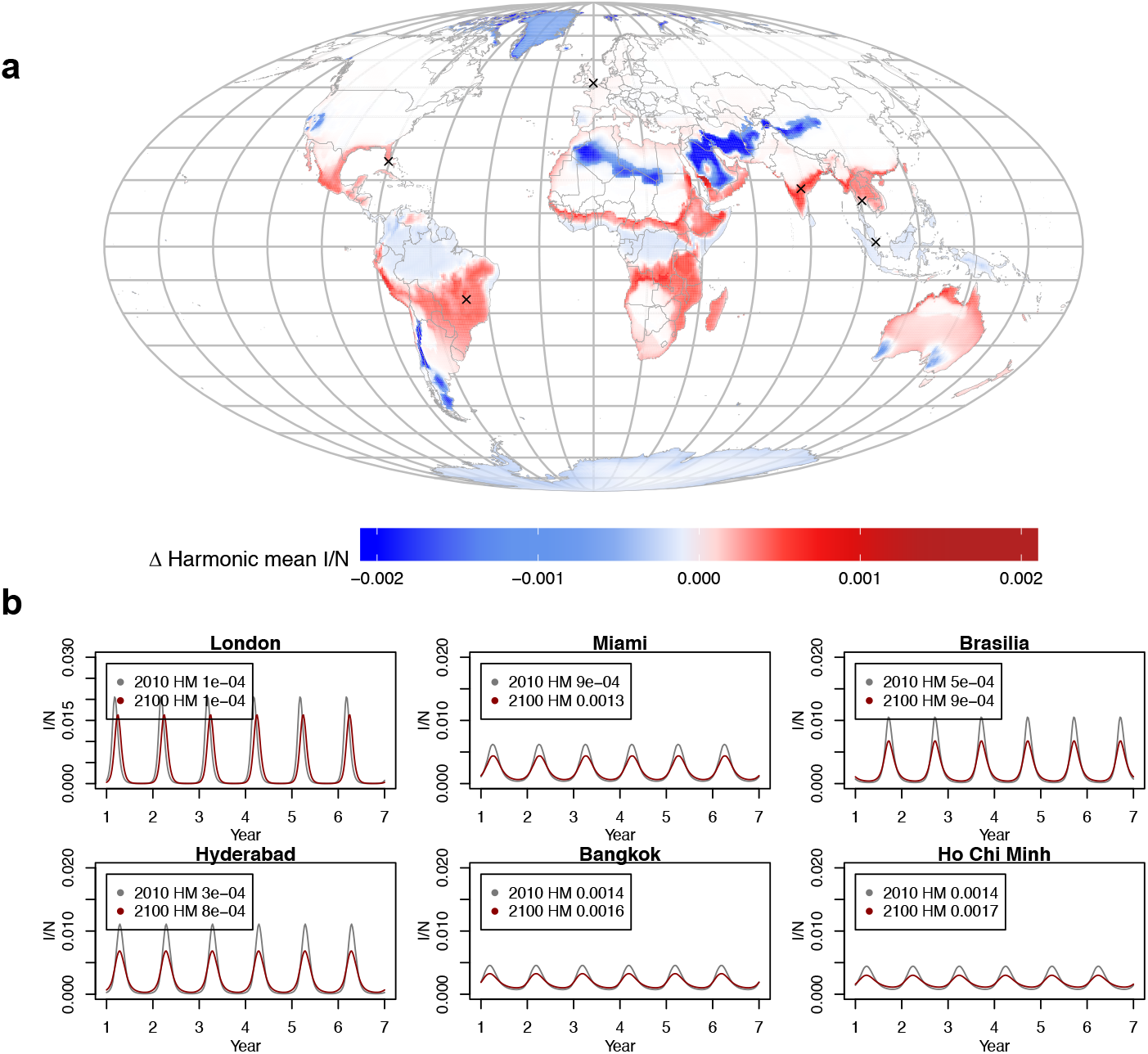
Climate and harmonic mean incidence (2010-2100). a) The effect of climate change on harmonic mean incidence per capita (HMI*), globally. Example locations for subfigure b are shown with “x”. b) Modelled incidence time series in 2010 versus 2100 for example cities representing a range of spatial locations and potential changes (additional examples shown in Fig. S4). HMI* is shown in the legend.

While Fig. 3a suggests that climate change will lead to an increased global area with suitable conditions for year-round influenza evolution, this does not take into account the spatial distributions of human populations, which are required to sustain the influenza epidemic. We consider the effect of population growth, in concert with climate change, on our measure of year-round influenza evolution suitability. To do this we multiply population numbers in each location by the present or projected HMI*. We note that our measure of nucleotide diversity in Fig. 2 is divided by the number of samples - total nucleotide diversity is not observed but is also expected to scale with population. We find that in 2010, the majority of locations with high HMI are located in Asia (Fig 4a). This supports earlier findings that influenza evolution in this region may seed global strains [27, 37]. However, when we consider future population growth, we find substantial increases to HMI occur in the African continent, such that this region becomes a locus of high genetic diversity (Fig. S7,S8). At the same time, we find that HMI in Asia will also continue to grow over the coming century.

**Figure 4:**
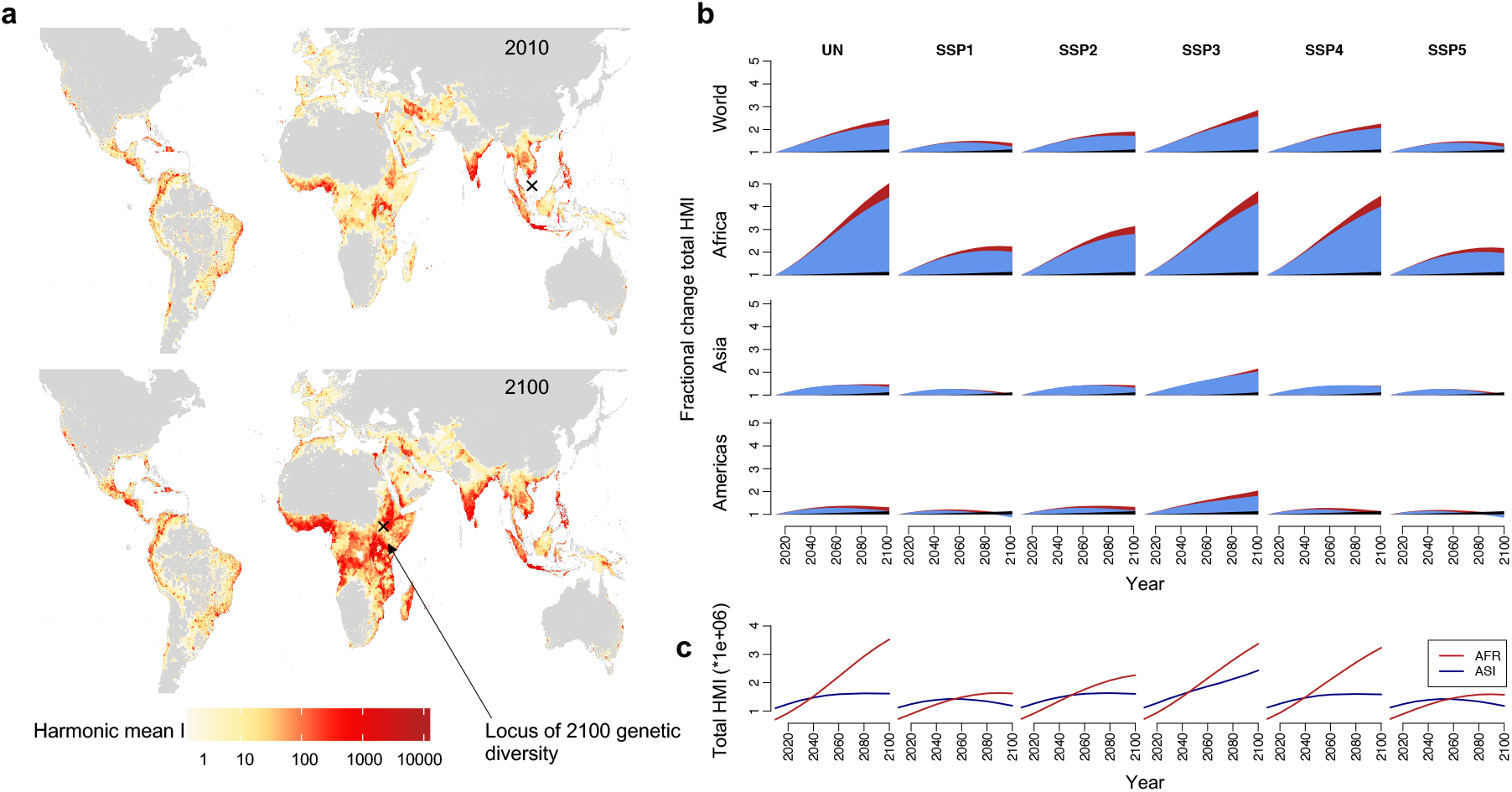
Population effects on harmonic mean incidence (2010-2100). a) Map of projected harmonic mean incidence in 2100 and 2010, based on CMIP5 RCP8.5 climate projections and UN population projections. “x” is the locus of genetic diversity in 2010 and 2100 (see Methods) b) The effect of climate (black), population (blue), combined (red) on fractional change in total harmonic mean incidence (number of pixels*HMI), relative to 2010 baseline. Different population growth scenarios are shown in columns. Different locations are shown in rows. c) The total harmonic mean incidence in African and Asia from 2010 to 2100. Model in all plots is run with *R*_*min*_ = 1.2 and *R*_*max*_ = 2.2.

Fig. 4b shows the percentage growth in total HMI relative to 2010. Here, we define total HMI as the sum of estimated HMI across all locations. We consider three scenarios: the effect of climate change alone, the effect of population growth alone and the combined effect. We also consider different population growth scenarios, as defined by the shared socioeconomic pathways, as well as UN projections for country-level population growth [38] (results using different Representative Concentration Pathways, shown in Fig. S5). Across all scenarios, the African continent sees a significant increase in HMI, driven primarily by population growth (Fig. 4b). In the most extreme scenario, the area with high HMI in Africa increases by a factor of 5 in 2100 relative to 2010. In all scenarios, Africa exceeds Asia as the primary location for influenza evolution at some point in the mid-21st century (Fig. 3c). Further, in all population scenarios, global suitability for evolution doubles in the coming century (first row Fig.4b).

Although population growth (blue slice in Fig. 4b) dominates the picture, the impact of climate change (black slice Fig. 4b) is not negligible. Without population growth, climate change leads to a net 13% increase in evolution suitability globally by 2100. The coupled effect of climate change and population growth lead to the largest changes in HMI surface area. Climate change leads to an additional 14% - 27% increase in HMI surface area by 2100 globally, depending on population growth scenario. For some countries, climate change plays an even bigger relative role. In the Americas climate change increases evolution suitability by 13%, with population changes alone leading to a comparable 14% change, in the UN growth scenario.

## Discussion

Our results imply that climate change will lead to more persistent outbreaks of influenza, with a average reduction in the size of seasonal peaks. We find that increased persistence, as measured by harmonic mean incidence, is correlated with year-round diversity of influenza strains and climate change may have subtle but important impact on increasing year-round strain diversity across locations. In contrast, we find that the rapid increase in global population projected over the coming century could have a substantial impact on influenza strain diversity. Higher population numbers located in increasingly dense urban locations in the tropics, where climate maintains year-round circulation of the virus, could lead to an increase in total strain diversity. In particular, we find that locus of diversity shifts from Asia in 2010 to Africa in 2100.

There are important caveats to these results. Primarily, the link between the size of population bottleneck and genetic diversity, or HMI and genetic diversity, remains controversial. Some studies assert that influenza evolution does not occur in a particular location, but happens as strains circulate globally [24]. A highly mobile population in Asia may contribute to current strain emergence in this region [27, 37]. We expect that even within this scenario, the existence of higher host numbers driven by population growth, as well as increased mobility, should heighten the risk of evolving novel strains. Urbanization may also lead to more persistent outbreaks, potentially affecting evolution [8]. However more work is needed to connect influenza evolutionary patterns, via phylogenetic analyses, with empirical data on climate and demographics. The impact of climate, as well as animal and human population distributions, on future pandemics is also an important question, albeit a difficult one to address.

Our climate change projections for influenza incidence support earlier work suggesting seasonal epidemics may become milder as the climate warms [39]. An obvious corollary is that we should observe a lower reproduction number at present in tropical locations, of which there is some evidence [40]. Interesting analogies may apply by considering the dynamics of influenza B, which has a lower *R*_0_ and has a polyphyletic tree for the surface protein (hemagglutinin), compared to the monophyletic tree for influenza A. Drilling into such comparative details is an important area for future work. While our results are based on changes to specific humidity, we find that including a hypothesized effect of precipitation on transmission does not alter our main conclusions (Fig. S9) [17, 41]. Our results suggest that while climatic and demographic change may lead to milder epidemics in the future, this could come at the cost of higher risk for evolved strains. Further, the African continent could become a locus for new strain emergence. This may be exacerbated by increased mobility in the region, as populations grow, a factor not explicitly addressed here. Increasing surveillance of influenza strains in Africa could help control efforts in the future. More broadly, our results indicate that while climate change will have a substantial impact on public health, one must also consider the projected major changes to population in the context of infectious agents.

## Methods

### Data

Historical specific humidity data come from NASA’s Modern-Era Retrospective analysis for Research and Applications (MERRA) dataset, available gridded at a resolution of 0.5°latitude and 0.625°longitude. Climate change projections come from the multi-model “business as usual” (RCP8.5) scenario of the Coupled Model Intercomparison Project Phase 5 (CMIP5), linearly downscaled to match the MERRA data, with specific humidity derived from temperature and relative humidity projections (holding relative humidity constant [42]). We average projected (and baseline) climate over a 10 year window preceding the target data i.e. 2100 climate is 2091-2100 averaged). Climate data is shown within land borders. The land border shapefile was downloaded from *thematicmapping*.*org*.

Baseline population data come from the Center for International Earth Science Information Network (CIESIN). Future projections are based on data from the United Nations World Population Prospects as well as the Global Population Projection Grids Based on Shared Socioeconomic Pathways (SSPs), accessed via CIESIN [38]. In the UN population scenario, we take the CIESIN gridded population data and multiply by the national-level UN projected changes. This assumes no change to spatial density of population. SSP population projections for 2010 and all following years are taken from the gridded datasets developed by Jones et al.[38] The UN projections and the SSPs have slightly different baselines.

### Model

Our influenza model is based on [10], specifically:

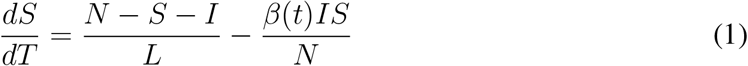

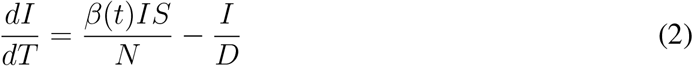

where *S* is the susceptible population, *I* is the number of infectious individuals and *N* is the population. *L* represents the duration of immunity and *D* is the mean infectious period. *β*(*t*) is the contact rate at time *t* and is related to the basic reproductive number by *R*_0_(*t*) = *β*(*t*)*D. R*_0_ has a dependency on specific humidity *q*(*t*), based on laboratory experiments [20, 10], given by:

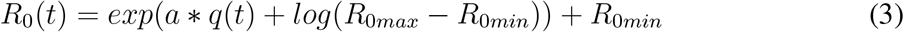

where *a* is found to be −180 and *R*_0*max*_, *R*_0*min*_ are the maximum and minimum daily reproductive numbers respectively.

Here, we take values for *R*_0*min*_ and *R*_0*max*_ from published estimates [10]. In baseline projections, we use *R*_0*min*_ = 1.2 and *R*_0*max*_ = 2.2 as candidate values, though our results are robust to using a range of values (Fig. S3). We take *D* to be four days, representing (approximate) mean estimates from the literature (studies using a similar model have found values of 2-3.75 and 4.43-5.26 [43]). We use a value of 45 weeks to represent the duration of immunity, *L*. While this is shorter than the single outbreak estimates used in earlier studies, this value allows epidemics to emerge annually, representing the evolution of novel strains to which hosts are not yet immune.

### HMI

Population genetics theory suggests that genetic diversity in a fluctuating population (in this case, virus population) may be related to the harmonic mean of the population size over time [29, 35, 34]. The harmonic mean provides a measure of the effective population size and has been applied in studies of virus populations [36], and more recently to epidemiological scenarios [33]. We assume population size is equal to incidence in hosts, measured at each generation time step (assumed to be a week). We first define harmonic mean incidence per capita as:

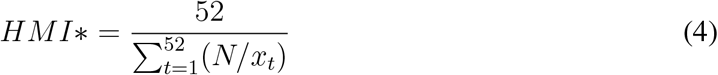

where *x*_*t*_ is incidence at time *t*, where *t* is a week in a year. *N* is the human population in a particular location. Because our model is deterministic, weekly incidence is the same year-to-year, so *HMI\** is dependent on intra-annual changes to incidence only.

*HMI\** gives the per capita harmonic mean incidence within a particular location, and allows us to consider global suitability for influenza evolution without accounting for demographic factors. We also define HMI by:

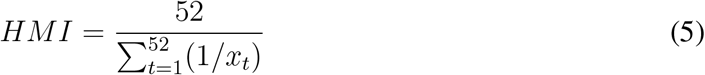

which is the absolute harmonic mean incidence, accounting for demography. In practice, we first calculate *HMI\** by running an influenza model for all locations, setting *N* = 1 and then multiply this by gridded population size numbers i.e.

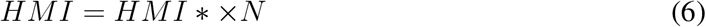

HMI is highly correlated with the absolute depth of trough, measured as the minimum annual incidence (Fig. S2).

### Nucleotide diversity

We analyzed HA sequence dataset of human influenza viruses H1NX and H3NX in tropical regions and north temperate regions, using sequences from the Global Initiative on Sharing All Influenza Data (GISAID). We chose six representative locations with sufficient data avalability at the sub-yearly level. We selected England and New York to represent north temperate locations, while Hawaii, Hong Kong, Singapore and Taiwan for tropical locations, with the sampling date from 2007 to 2019 (a time period in which all locations had data). Only sequences covering at least 70% of full length were retained. Sequences were annotated with available collection date and aligned with MAFFT [44], and edited manually. Accession numbers of sequences are provided at *github/rebaker64/flu*.

With the sequence data, we calculated the pairwise nucleotide distance between sequences under the molecular evolutionary models of K80 [45] with ‘ape’ package [46] in R v3.5.2. To get the nucleotide diversity in each quarter of each year, we divided the sum of pairwise distances between sequences in each quarter by the number of comparisons [47]. With the quarterly nucleotide diversity, we averaged the diversity by quarter over 13 years (2007 – 2019). If no sequences were available for a particular quarter and location, the diversity was recorded as zero.

### Locus of genetic diversity

We calculate the inverse distance weighted average of HMI for each location on the globe (using minimum distances on the spherical surface). The location with the maximum average value is labelled as the locus of genetic diversity. Locations with the top 10% HMI in 2010 and 2100 are shown in Fig. S7.

## Data Availability

Data is publicly available. Code available at https://github.com/rebaker64/flu.

## Acknowledgements

This work is supported by the Cooperative Institute for Modeling the Earth System (CIMES) (REB), US National Institutes of Health grant GM110748 (JS), Natural Sciences and Engineering Research Council (NSERC) of Canada (PGS-D to CMSR), the James S. McDonnell Foundation 21st Century Science Initiative Collaborative Award in Understanding Dynamic and Multi-scale Systems (CMSR).

## Author Contributions

Conceptualization: REB, GV, CJEM, BTG; Data curation: REB, GV, WY; Formal analysis: REB; Methodology: REB, CJW, CMSR, CV, JS, CJEM, GV, BTG; Software: REB; Visualization: REB; Writing, original draft: REB, BTG; Writing, reviewing and editing: REB, CJW, CMSR, CV, JS, CJEM, GV, BTG.

## Data Availability

All data used in this study are publicly available. Baseline climate data are available from NASA’s Modern-Era Retrospective analysis for Research and Applications (MERRA). Climate projection data is downloaded from KNMI Climate Explorer. Baseline population data are from the Socioeconomic and Data Applications Center and SSP population projections are from the same source. We also use population projections from the UN Department of Social and Economic Affairs. Hong Kong influenza data used in Fig. S9 is available from the Department of Health, The Government of the Hong Kong Special Administrative Region. The complete accession list of sequences used in the phylogenetic analysis are available at *github/rebaker64/flu*. Sequence data come from the Global Initiative on Sharing All Influenza Data (GISAID).

## Code Availability

Code for recreating the main results is available at *github/rebaker64/flu*.

