## Supplementary Information for "Implications of climatic and demographic change for seasonal influenza dynamics and evolution"

#### Contents

**1 Supplementary Figures** **2**

### 1 Supplementary Figures

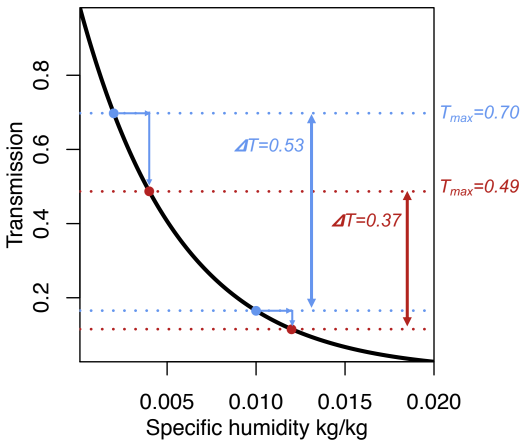


Figure S1: **Schematic of transmission and climate dependence**. Transmission is related to specific humidity non-linearly. A hypothetical uniform annual increase in humidity (blue horizontal arrows) results in a decline to maximum transmission (blue = baseline, red = with climate change) as well as the range of transmission values a location experiences throughout the year (vertical arrows).

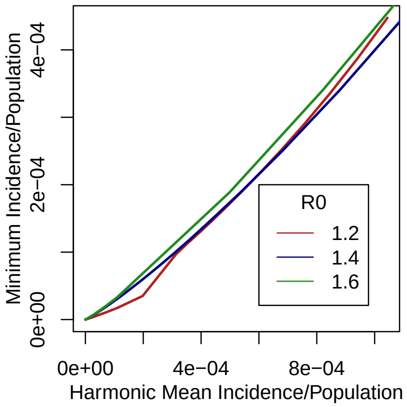


Figure S2: **The correlation between harmonic mean incidence and minimum cases**. There is a close relationship between HMI* (per capita) and minimum cases, for a hypothetical population under different scenarios of baseline $R_0$.

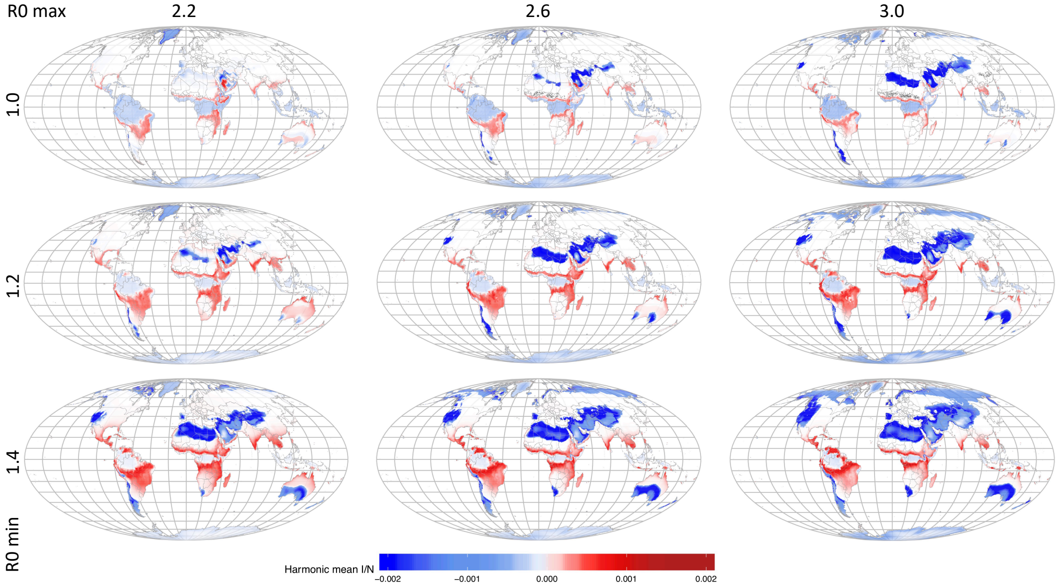


Figure S3: **Uncertainty in harmonic mean incidence projections**. The effect of climate change on HMI* (2100 - 2010) for different $R_0$ values. In the main model, $R_0 min$ = 1.2 and $R_0 max$ = 2.2 (shown in Fig. 3).

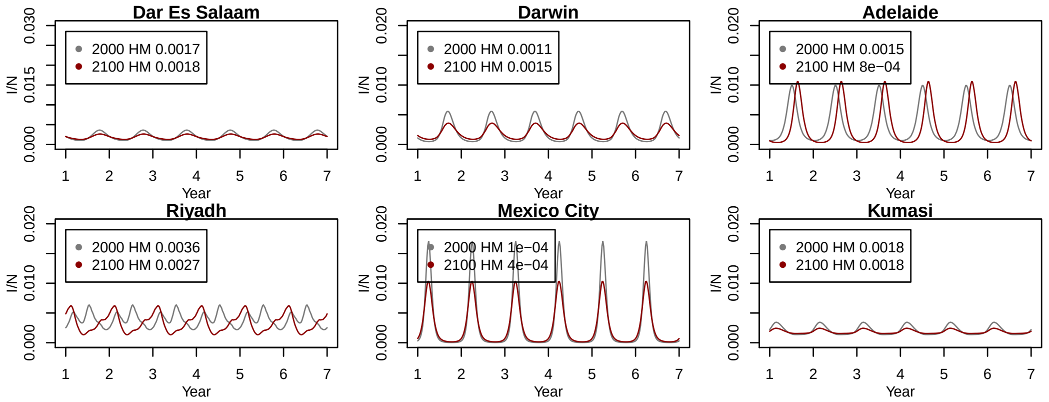


Figure S4: **Example model predictions**. The effect of climate change on future influenza dynamics for different example cities. Locations chosen to reflect a range of projected changes and spatial locations.

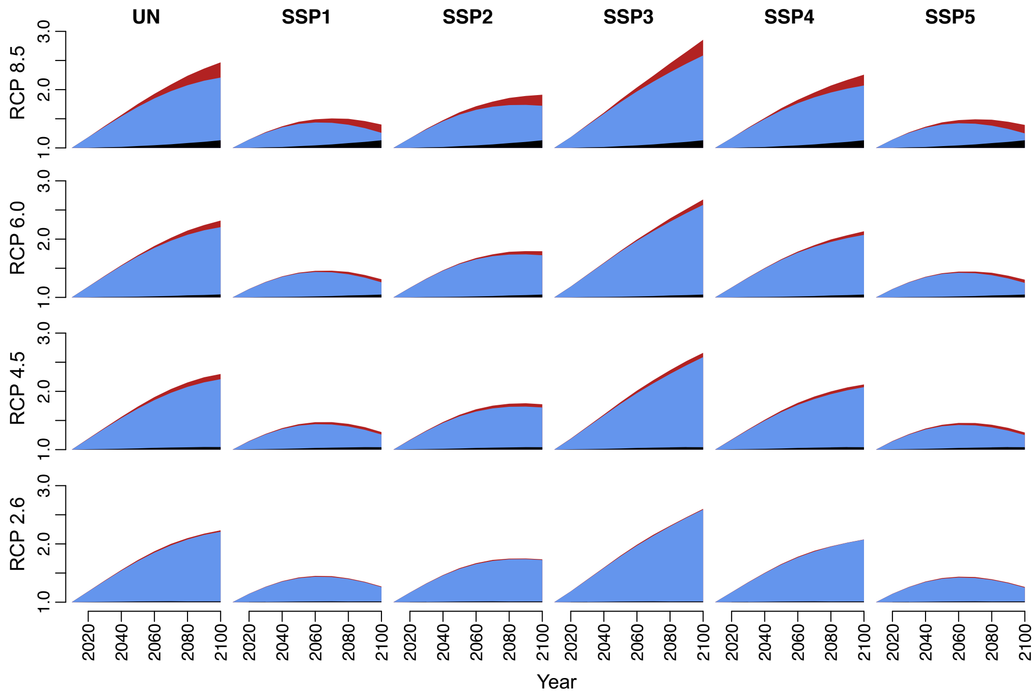


Figure S5: **Future changes in harmonic mean incidence (HMI) by Representative Concentration Pathway (RCP)**. The effect of population growth (light blue), climate change (black) and both combined (red) on the total change in HMI, relative to a 2010 baseline. The population growth effect dominates in all scenarios.

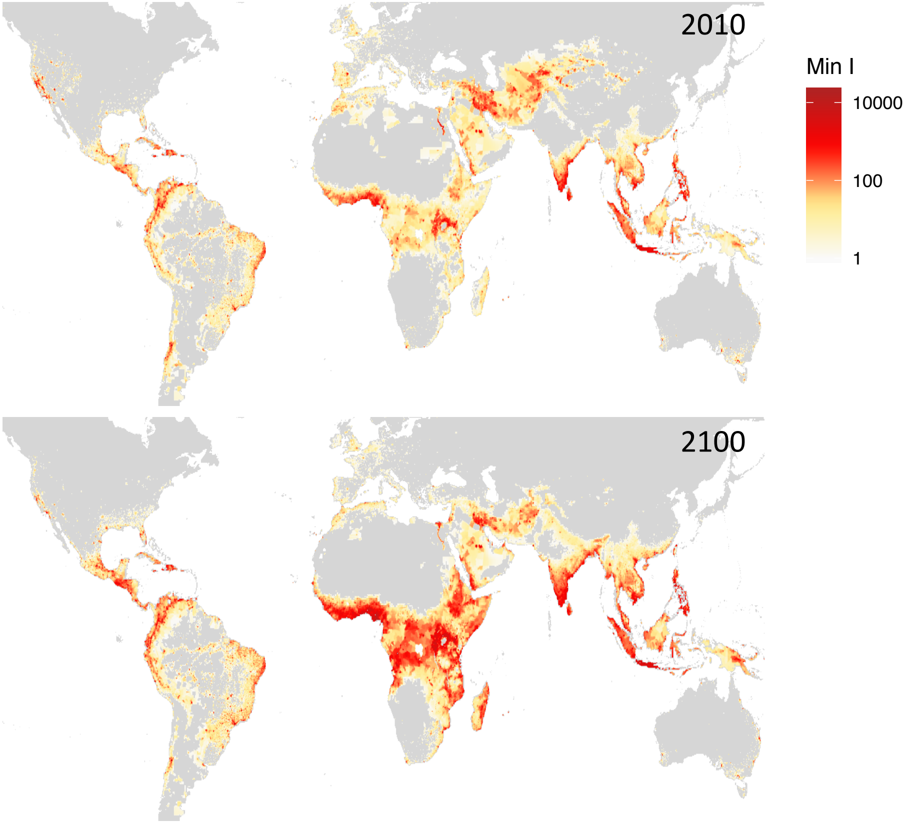


Figure S6: **Minimum incidence 2010 and 2100.** Map showing annual minimum influenza incidence in 2010 and 2100. This represents the depth of the incidence trough and has a high correlation with harmonic mean incidence (for comparison, see Fig. 4a).

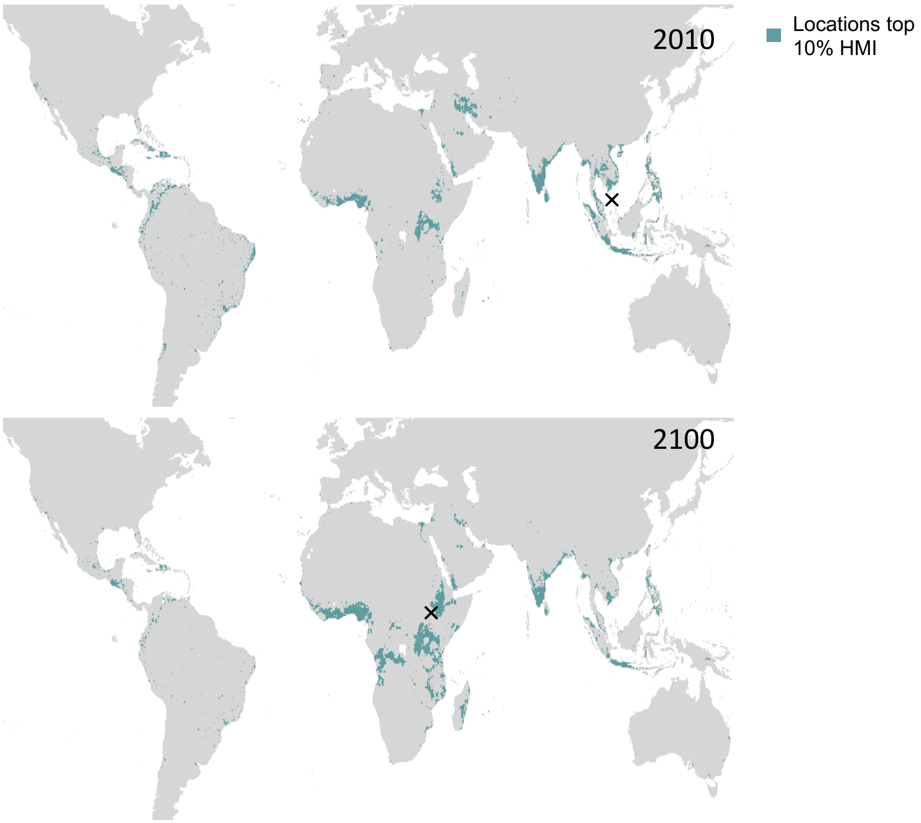


Figure S7: **Top 10% HMI locations 2010 and 2100.** Map showing locations with the 10% highest Harmonic Mean Incidence in 2010 and 2100. While dispersed across the tropics, most locations are in Asia in 2010 and Africa in 2100.

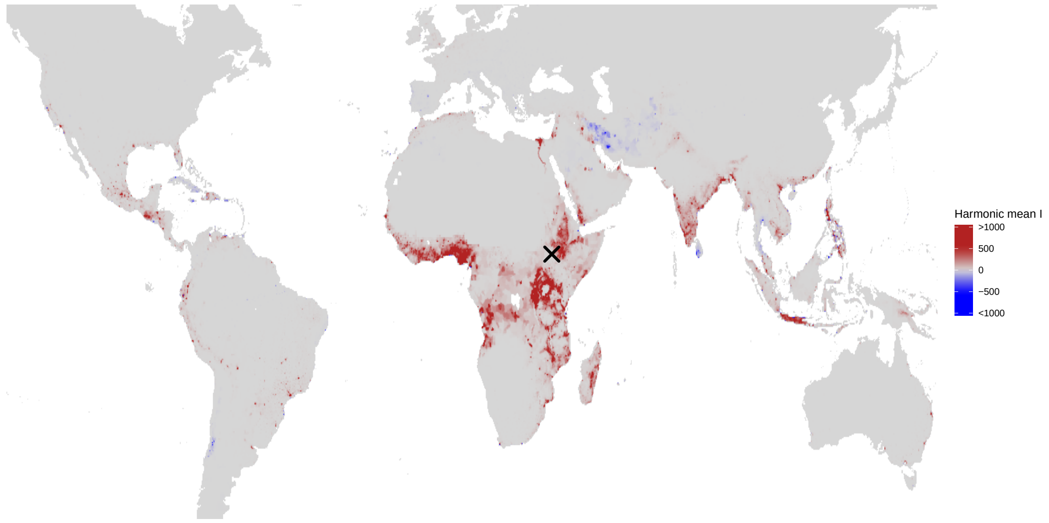


Figure S8: **Change in HMI 2100-2010.** Change in HMI between 2100 and 2010 using UN projected values for population and RCP8.5 climate projections. This is the difference between the two maps in Fig. 4a.

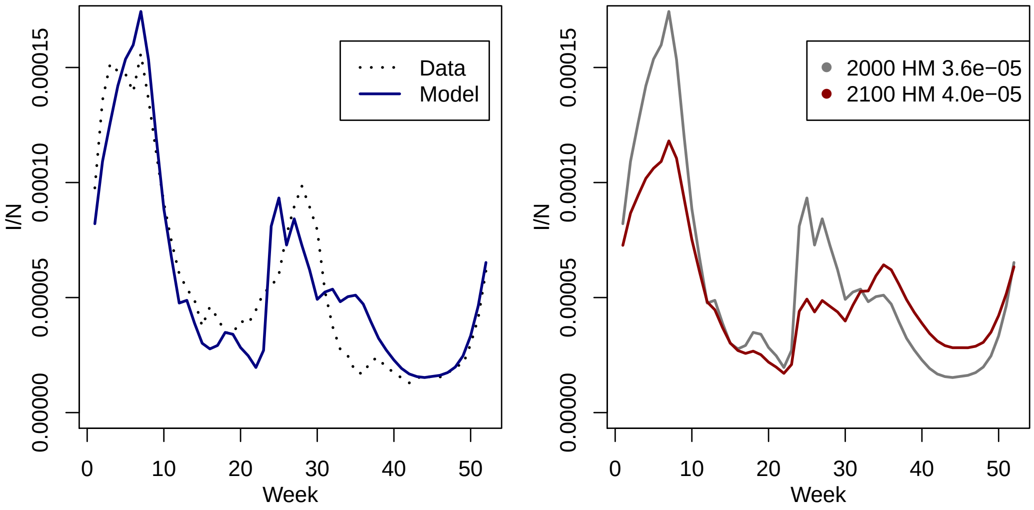


Figure S9: **Influenza model fit for Hong Kong.** We fit our influenza model to data from Hong Kong, with the fitted parameters being the precipitation effect on transmission and a constant reporting rate. The left-hand figure shows the model fit: the model is able to capture the seasonality of Hong Kong influenza cases. The right-hand figure shows the effect of projected precipitation and humidity changes on HMI (not accounting for under-reporting).
